# Projected economic gains and lives saved under universal healthcare in the United States

**DOI:** 10.64898/2026.07.22.26358689

**Authors:** Abhishek Pandey, Chad R. Wells, Yang Ye, Meagan Fitzpatrick, Alison P. Galvani

**Affiliations:** Center for Infectious Disease Modeling and Analysis (CIDMA), Yale School of Public Health, New Haven, CT, USA; Center for Vaccine Development and Global Health, University of Maryland School of Medicine, Baltimore, MD, USA

## Abstract

The US spends more on health care than any other nation, yet tens of millions of Americans are uninsured or underinsured, and coverage retractions enacted in 2025 are widening these gaps. The misalignment between the for-profit insurance architecture and optimal patient care, together with the inefficiencies of a fragmented system, contributes to both unnecessary costs and preventable mortality. We update our previous analyses with the most recent data to project the economic benefits and the number of lives saved that would be achieved by single-payer universal coverage, as proposed in the Medicare for All Act. We estimate that such a system would reduce national health expenditure by $1,041 billion annually. Sources of savings include reductions in administrative overhead, pharmaceutical prices, fraudulent billing, and avoidable emergency care. Combined with the reversal of recent retractions, universal coverage would save over 114,000 lives annually.

## Introduction

The United States spends more on health care than any other nation, an estimated US$5.3 trillion in 2024, equivalent to nearly one fifth of gross domestic product.^1^ Yet a greater share of the US population is exposed to the financial and clinical consequences of inadequate coverage than in comparable high-income countries. In 2024, approximately 27.5 million Americans lacked health insurance,^2^ while tens of millions more were underinsured, struggling under deductibles and cost-sharing requirements that placed necessary care beyond financial reach.^3^ Despite its extraordinary level of expenditure, the United States continues to trail its peers in life expectancy, preventable mortality, and other major measures of population health.^4,5^ Exceptionally high spending, incomplete coverage, and comparatively poor health outcomes therefore remain defining features of the US health-care system and reflect the substantial human and economic toll of maintaining the status quo.

That toll is rising as recent coverage gains are being reversed. The current US system relies on a patchwork of subsidies to facilitate enrollment by seniors and low-income people into insurance plans that they could otherwise not afford. These subsidies have been rolled back or blocked by a series of Congressional decisions in 2025 and 2026, with approximately five million people anticipated to lose coverage and many seniors moving from fully insured to underinsured.^6,7,8,9^ After a decade in which uninsurance declined, the gap between what Americans pay for health care and what they receive in return is widening once again.

Against this backdrop, single-payer universal coverage, as proposed in the Medicare for All Act,^10^ remains the most comprehensive alternative to the current system. For six decades, Medicare has provided coverage to Americans aged 65 years and older. Extending a similar public insurance structure to the entire population would replace the fragmented for-profit multipayer system with a single national program. Since our previous analysis of Medicare for All published in 2020,^11^ mounting healthcare system costs have continued to outpace the growth of our GDP. In light of this trajectory, the changing policy landscape, and an aging population, we update our analyses to incorporate the most recent data and new status quo for the healthcare system.

In this study, we use the most recent National Health Expenditure data and current insurance coverage estimates to assess the system-wide economic and mortality consequences of transitioning to a single-payer universal healthcare system. We estimate changes in national health expenditure by sequentially accounting for the application of Medicare payment rates to all providers, pharmaceutical pricing under international reference pricing, administrative simplification, reductions in fraud, and the avoidance of emergency and inpatient care that timely access to primary care would make unnecessary, alongside the additional spending that universal coverage would entail: the reimbursement of care that currently goes unpaid, the greater use of health services by people who are currently uninsured or underinsured, and the extension to include dental coverage. We then estimate the associated mortality effects using an attributable-fraction framework that incorporates the elevated mortality risks of both uninsured and underinsured populations.

## Methods

We modeled the transition from the current multi-payer system to single-payer universal coverage and estimated two outcomes: the change in national health expenditure and the change in annual mortality, both for the 2024 reference year. Baseline spending by type of service and source of funds was taken from the National Health Expenditure Accounts^1^; insurance coverage by age from the American Community Survey^2^; the prevalence of underinsurance from the Commonwealth Fund Biennial Health Insurance Survey^3^; and age-specific deaths from national vital statistics.^12^ Full definitions, parameter derivations, and calculations are provided in the **Supplementary Appendix**.

### Health-expenditure model

We estimated national health expenditure under universal health care by applying a sequence of adjustments to the 2024 baseline, following a previously described stepwise accounting framework^11^ updated with 2024 data and extended with a dental-coverage step. We first separated the expenditures of the Department of Veterans Affairs, the Department of Defense, and the Indian Health Service, which would continue to operate in parallel to Medicare, and consolidated pharmaceutical spending into a single category. From this baseline, each subsequent step raises or lowers recognized national spending through a distinct mechanism (**Table 1)**.

**Table 1:** Stepwise change in US national health expenditure under a single-payer universal health care system, 2024. Values in US$ billions. Full parameter derivations and sources are given in the **Supplementary Appendix**.

| Step | Adjustment | Change | Cumulative |
| --- | --- | --- | --- |
|  | **Baseline national health expenditure, 2024.** Total US health spending by type of service and source of funds.<sup>1</sup> |  | 5278.6 |
| 1 | **Separate parallel federal systems; consolidate pharmaceutical spending.** Spending by the Department of Defense, the Department of Veterans Affairs, and the Indian Health Service is separated. Pharmaceuticals administered as part of service provision are consolidated into a single category. | 0.0 | 5278.6 |
| 2 | **Recognize previously uncompensated care.** Hospital care currently delivered but never reimbursed is added to recognized spending, since a universal system would pay for all legitimate care. | +51.7 | 5330.2 |
| 3 | **Avert avoidable emergency and inpatient use.** Emergency-department visits and hospitalizations that would be prevented by timely access to primary care are removed. | −100.0 | 5230.2 |
| 4 | **Pay all providers at Medicare rates.** All payers converge on Medicare rates, applied separately to hospital and to physician and clinical services. | −295.6 | 4934.7 |
| 5 | **International reference pricing on pharmaceuticals.** Prices are benchmarked to those paid in comparable high-income countries, yielding a 51% reduction in pharmaceutical spending.<sup>13</sup> | −377.5 | 4557.1 |
| 6 | **Reduce administrative overhead.** Consolidating billing and insurance functions into a single payer reduces overhead down to the level of the current Medicare program.<sup>19</sup> | −286.3 | 4270.8 |
| 7 | **Reduce fraudulent billing.** A unified claims system enables detection of improper billing, applied as an 8% reduction, consistent with single-payer transitions elsewhere.<sup>14</sup> | −285.7 | 3985.1 |
| 8 | **Coverage-induced utilization.** People who are currently uninsured or underinsured use less care than the adequately insured because of cost barriers; their use is raised to the adequately-insured level.<sup>11,20</sup> | +197.7 | 4182.7 |
| 9 | **Universal dental coverage.** Dental service use is raised toward the level observed among the privately insured.<sup>15</sup> | +54.7 | 4237.4 |

Four adjustments reduce national expenditure. Hospital and clinical fees are paid at Medicare reimbursement rates, which are below the rates paid by commercial insurers and above those paid by Medicaid. Pharmaceutical prices are reduced through international reference pricing, applying the 51% reduction^13^ achieved by benchmarking US prices to those paid in comparable high-income countries. Administrative overhead is reduced toward the level of the current Medicare program, reflecting simplification of billing and insurance functions under a single payer. Fraudulent billing is reduced, consistent with the experience of other single-payer transitions.^14^ A further reduction comes from averting emergency-department visits and hospitalizations that improved primary-care access would render unnecessary.

Three adjustments increase national expenditure, reflecting care that is currently unmet or unpaid. Previously uncompensated care, principally unreimbursed hospital care, is added to recognized spending, since a universal system would reimburse all legitimate care. The use of services by people who are currently uninsured or underinsured is raised to that of the adequately insured, who face fewer cost barriers. Finally, universal dental coverage is represented as an increase in dental use toward the level observed among the privately insured.^15^

### Mortality model

We estimated the mortality effect of universal coverage using an attributable-fraction framework, in which the observed deaths in each age group arise from the current mix of insured, uninsured, and underinsured individuals of that age, with mortality rates dependent on insurance status.^11^ We then calculated the deaths that would be expected if the entire population were adequately insured.

For the uninsured, we assigned a mortality hazard ratio of λ = 1.40 relative to the adequately insured, consistent with cohort estimates of excess mortality among uninsured adults.^16^ We then extended this framework to the underinsured, whose excess mortality has not previously been incorporated. Direct mortality estimates for the underinsured are not available, so we derived a baseline hazard ratio of γ = 1.25 by interpolating between the adequately insured and the uninsured according to the relative prevalence of cost-related forgone care in each stratum, the mechanism through which inadequate coverage is thought to raise mortality (**Supplementary Appendix**).^3^ Underinsurance was assigned to working-age adults (19–64 years), among whom approximately 23% are underinsured,^3^ and set to zero at ages 0–18 and 65 and over, where near-universal public coverage makes underinsurance uncommon. Applying the age-specific uninsured and underinsured shares and their hazard ratios to observed deaths,^12^ we estimated the annual lives saved by universal coverage. Given that coverage losses as well as other retractions^8^ now underway have largely materialized in the period since the latest available data (2024), and would be reversed under a universal system, we additionally accounted for the further lives saved by reversing these retractions, drawing on our previous published projections^17^ to report the overall annual lives expected from enactment of a universal health care.

### Sensitivity analyses

We assessed the robustness of both health and economic outcomes to their principal parameters. For expenditure, we varied the pharmaceutical price reduction. The scenarios we considered were international reference pricing, the 23.1% discount negotiated by the Department of Veterans Affairs,^13^ and no reduction. We considered a scenario without fraud adjustment. For mortality, we varied the underinsured hazard ratio γ across its plausible range from 1.00 to 1.40, where γ = 1.00 attributes no excess mortality to underinsurance and recovers the uninsured-only estimate; we report the resulting range alongside the γ = 1.25 baseline.

## Results

### System-wide savings

US national health expenditure in 2024 was US$5278.6 billion. Under a single-payer universal health care system, we estimate that national health expenditure would become US$4237.4 billion, a reduction of US$1041.2 billion, or 19.7% of current spending (**Table 1**).

The net savings are the balance of two opposing sets of effects (**Figure 1)**. Reductions total US$1345.2 billion, driven primarily by international reference pricing on pharmaceuticals (US$377.5 billion), application of Medicare payment rates to all providers (US$295.6 billion), reduced administrative overhead (US$286.3 billion), and reduced fraudulent billing (US$285.7 billion), with a further US$100.0 billion from averting avoidable emergency and inpatient care. No single mechanism accounts for more than approximately one quarter of the gross reduction. Savings are partly offset by US$304.0 billion of additional spending, comprising the expanded use of services by people who are currently uninsured or underinsured (US$197.7 billion), universal dental coverage (US$54.7 billion), and the compensation for care that is currently delivered but never reimbursed (US$51.7 billion). Expanded access therefore consumes approximately one fifth of the gross reduction, and the system remains substantially cost-saving even after it is fully accounted for.

**Figure 1:**
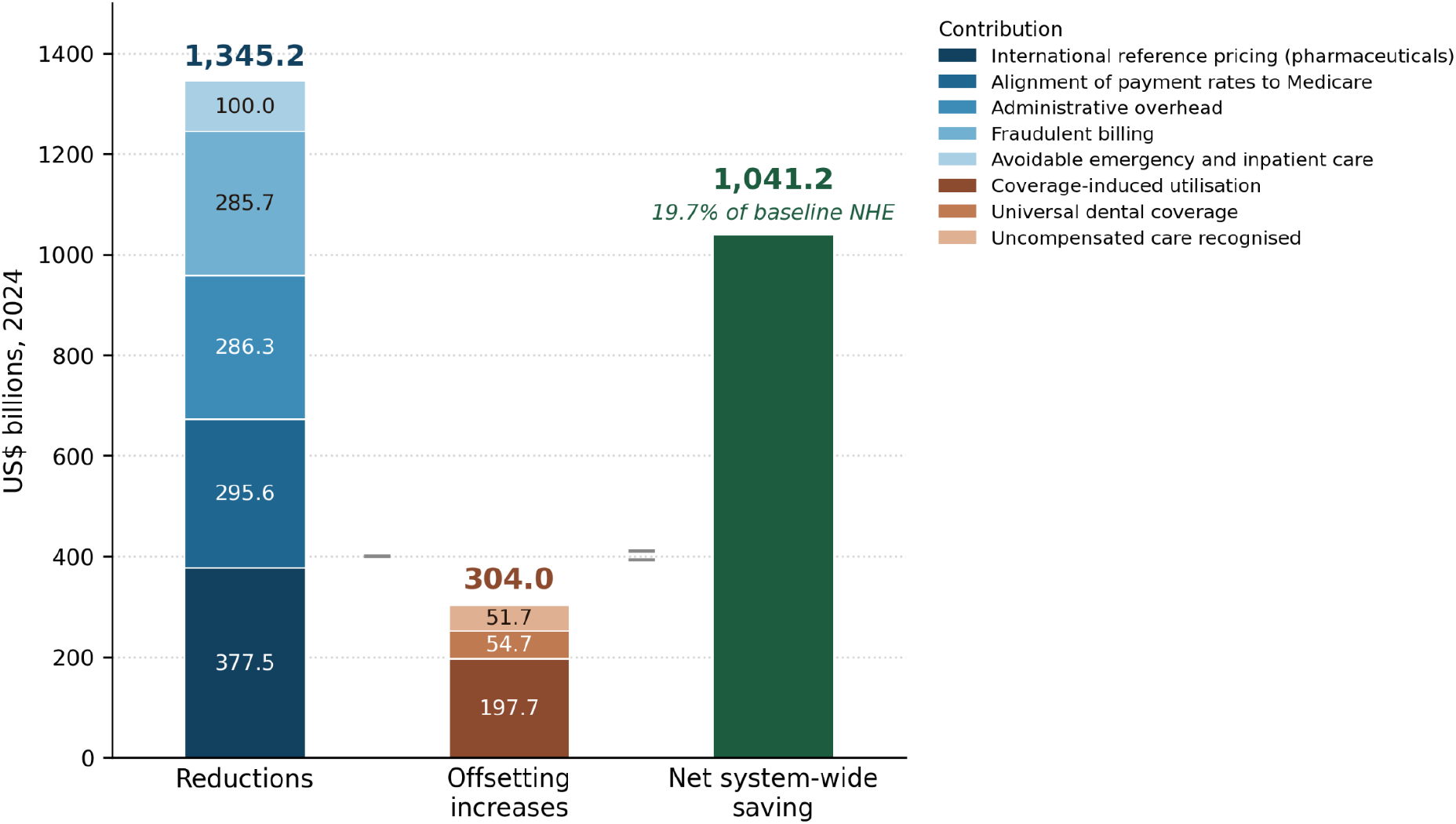
Composition of the system-wide saving under a single-payer universal health care system, United States, 2024. Five mechanisms reduce national health expenditure by US$1345.2 billion, and three increase it by US$304.0 billion, giving a net saving of US$1041.2 billion, or 19.7% of the 2024 baseline of US$5278.6 billion. Parameter values and sources are given in **Table 1** and **Supplementary Appendix**.

Savings are robust even with adjustments in the parameters for which the underlying evidence is least direct (**Table S3**). Substituting pharmaceutical prices consistent with those negotiated by the Department of Veterans Affairs, removing the pharmaceutical price reduction altogether, and omitting the savings from fraud detection, each lower the estimate, but under every variation considered the system-wide saving remains at least US$663.3 billion, or 12.6% of current spending.

### Lives saved

In 2024, 27.5 million people in the USA had no health insurance, and more than 45 million adults aged 19–64 years were underinsured. Applying a mortality hazard ratio of 1.40 for the uninsured and 1.25 for the underinsured, we estimate that universal adequate coverage would save 62,863 lives annually relative to the current system (**Figure 2**).

**Figure 2:**
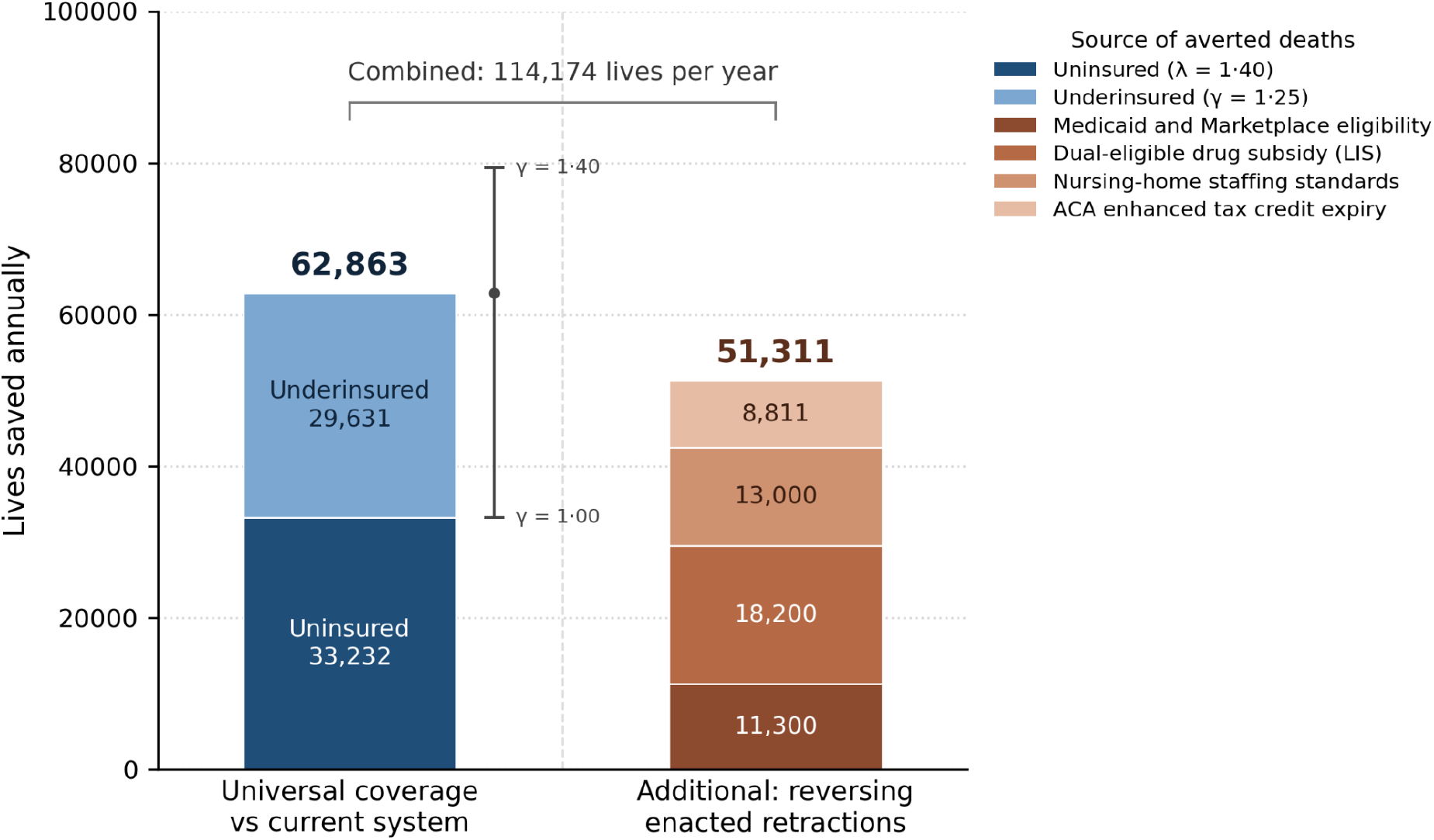
Annual lives saved under a single-payer universal health care system, US, 2024. The left bar shows deaths averted each year by extending adequate insurance to the entire population relative to the current system, decomposed into deaths attributable to uninsurance (hazard ratio λ = 1.40 relative to the adequately insured, held fixed throughout) and to underinsurance (γ = 1.25, baseline). The whisker shows the sensitivity of this total to the underinsured hazard ratio across its plausible range (γ = 1.00 to 1.40); at γ = 1.00 no excess mortality is attributed to underinsurance and the estimate reduces to the uninsured-only case. The right bar shows the additional deaths a universal system would avert by reversing coverage retractions enacted since 2025.^17^

Approximately half of this total is attributable to each stratum: 33,232 prevented deaths (52.9%) arise from the currently uninsured and 29,631 (47.1%) from the currently underinsured. The underinsured therefore account for a mortality burden comparable in magnitude to that of the uninsured, despite holding insurance.

The estimate is sensitive to the hazard ratio assigned to the underinsured. Across the plausible range of this hazard ratio γ from 1.00 to 1.40, annual lives saved would range from 33,232 to 79,367 (**Figure 2**). The lower bound corresponds to the assumption that underinsurance results in no excess mortality, and the upper bound to the assumption that underinsurance has the same impact on mortality as uninsurance.

### Additional lives saved by reversing coverage retractions

Beyond the deaths averted relative to the current system, a universal system would reverse coverage retractions enacted in 2025, which have been projected to cause 51,311 additional deaths annually (**Figure 2**).^17^ The largest share, 20,111 deaths, arises from people losing insurance altogether: 11,300 from restrictions on eligibility for Medicaid, including work requirements,^18^ and for subsidised private coverage, and 8,811 from the expiry of the enhanced subsidies for private coverage. A further 18,200 deaths follow from the withdrawal of assistance that helps low-income elderly people meet the cost of prescription drugs, and the remaining 13,000 stem from the suspension of minimum staffing standards in nursing homes. Taken together with the 62,863 deaths averted relative to the current system, universal coverage would avert 114,174 deaths annually relative to its current trajectory.

## Discussion

We estimate that a single-payer universal health care system would reduce US national health expenditure by US$1041.2 billion annually, or 19.7% of current spending, while saving 62,863 lives each year relative to the current system. Accounting for coverage retractions enacted since 2025, which such a system would reverse, the annual mortality benefit rises to 114,174 deaths averted. Our findings indicate that the principal barrier to universal coverage in the USA is not the absence of resources, but their allocation.

Our estimates differ from our earlier work in this framework^11^ for several identifiable reasons. National health expenditure has grown substantially since 2017, enlarging the base to which each adjustment applies. Our estimate of the gap between commercial and Medicare payment rates is wider, reflecting both improved benchmarking of commercial prices and a genuine widening of that gap over the intervening period. We benchmark pharmaceutical prices to those paid in comparable high-income countries rather than to the formulary of the Department of Veterans Affairs, a mechanism that has become more directly relevant since Medicare acquired limited negotiating authority.^13,21^ Moreover, we incorporate the mortality implications of the underinsured, and we model dental coverage. The number of people without insurance has fallen from approximately 38 million to 27.5 million,^2^ so the deaths attributable to uninsurance alone are now fewer than previously estimated. This reflects the coverage gains of the intervening decade rather than any diminution of the case for universal coverage. However, upon incorporating mortality burden attributable to the underinsured as well as from retractions from the recent legislation,^8^ our total mortality burden exceeds earlier estimates.

We found that the underinsured account for nearly half of the avertable mortality we estimate, 29,631 of 62,863 deaths. These are people who hold insurance and who are ostensibly covered, yet whose deductibles and cost-sharing lead them to forgo care that would have prolonged their lives. The composition of the problem has shifted: uninsurance has declined since 2010, while underinsurance has risen.^3^ Policy and analysis that attend only to the uninsured therefore address a diminishing share of the mortality attributable to inadequate coverage. The objective is not coverage as such, but coverage adequate to remove cost as a barrier to care. This is what distinguishes single-payer universal coverage from incremental expansion: because the Medicare for All Act eliminates deductibles and cost-sharing rather than extending insurance that retains them, it addresses both components of this burden, whereas reforms that reduce uninsurance alone would leave a large and growing part of it in place.

Dental care is among the largest coverage gaps in the US health system: more than one in five adults aged 19–64 years has no dental coverage, roughly twice the proportion who lack medical insurance, and Medicare has never covered routine dental care.^15,22^ Access is correspondingly rationed by cost. In 2022, 53.1% of adults with private dental coverage had a dental visit, compared with 23.9% of those with public coverage and 15.2% of those with none.^15^ Extending coverage to the whole population, at the rate of use observed among the privately insured, would raise national dental spending by an estimated US$54.7 billion. Dental care under a universal system is therefore an expansion of access rather than a source of savings, and the system absorbs its cost comfortably. Both the Medicare for All Act and legislation introduced separately to add dental, hearing, and vision benefits to Medicare would cover these services,^10,22^ reflecting a recognition that the exclusion of dental care from Medicare is a gap rather than a boundary.

While we model universal dental coverage by raising utilization to the level observed among the privately insured, we do not extend the same treatment to vision and hearing, which the national health expenditure accounts do not report as separate categories and which therefore cannot be isolated in the same way. Both are covered under the Medicare for All Act,^10,22^ and their inclusion would add further access-driven cost. However, given that spending on each is a small fraction of dental spending, we expect this to be modest.

On the expenditure side, our analysis is a static accounting for a single year. It does not incorporate the costs of transition, the contraction of administrative employment that consolidation implies, or the responses of providers to Medicare payment rates, each of which would affect the timing though not the direction of the savings we project. We assume that people who are currently uninsured or underinsured would use services at the rate of the adequately insured, which determines the US$197.7 billion attributed to expanded use; if their unmet need is greater than this, as the higher prevalence of undiagnosed conditions among the uninsured suggests, the cost of coverage expansion would be larger.

A fragmented payment system is permissive of improper billing by construction: no payer observes a provider’s full claim history, so patterns that would be anomalous in aggregate, such as duplicate claims or implausible volumes of billed physician time, are dispersed across many insurers, each of which sees only a fraction of them. The loss to fraud in the US is estimated between 3–10% of total health expenditure.^23,24^ A single payer processing all claims through one system, under one set of coding rules and one audit standard, removes much of the opacity on which improper billing depends. Our estimate assumes that consolidation would recover a substantial share of what fragmentation currently conceals. Extrapolating from the reduction observed following Taiwan’s single-payer transition,^14^ we apply an 8% reduction to the categories in scope, equivalent to 5.4% of national health expenditure and therefore within the range attributed to fraud in the US.

Several features of our approach make our estimates conservative. We reduce the administrative overhead of insurers, but do not count the corresponding reduction in the billing burden borne by providers, which a unified payment system would substantially relieve and which prior work has valued in the hundreds of billions of dollars annually.^25^ Given our accounting is confined to a single year, it excludes the savings that follow from earlier diagnosis and preventive care, which accrue over a longer horizon. On the mortality side, we attribute excess mortality to underinsurance only among adults aged 19–64 years, although cost-sharing under Medicare leaves many older adults exposed to similar barriers^26^; since more than three quarters of deaths occur after age 65, this exclusion is substantial. We also count no benefit from the improvements in continuity of care that a single system would allow, nor from the reductions in morbidity that would accompany the mortality benefit we estimate. Our figures should therefore be read as a lower bound in these respects.

A universal health-care system in the US would not require the country to spend more on health care. It would require it to spend less. Our results indicate that the existing budget is more than sufficient to cover everyone at lower total cost. The obstacle to universal coverage has never been the absence of resources, but the way they are allocated. A system that covers everyone, costs $1 trillion less, and averts more than 100,000 deaths annually requires no new discovery to implement, only enactment.

## Data Availability

All data produced in the present work are contained in the manuscript.

