## Supplementary Appendix for "Projected economic gains and lives saved under universal healthcare in the United States"

We estimate the system-wide economic and mortality consequences of a single-payer universal health care system using the 2024 release of the National Health Expenditure (NHE) accounts.[1] The total 2024 US health expenditure (denoted $\Sigma t,0$ throughout) is $5,278.55 billion. We report two outcomes:

1. System-wide change in national healthcare spending (ΔNHE): the difference between baseline NHE and post-transition NHE, capturing the net change in total US healthcare spending.

2. Annual lives saved: the reduction in deaths that would follow from providing adequate insurance to the entire population, relative to the current system.

### 1. National Health Expenditure transformation

Following our previously developed approach,[2] we model the change in national healthcare spending through a stepwise pipeline that transforms the 2024 baseline NHE into post-transition NHE. Each step modifies an aspect of healthcare delivery, payment, or utilization that we expect to change under universal coverage. We extend the previously described eight-step pipeline with a ninth step representing universal dental coverage (Section 1.9).

We use the notation $\Sigma t,k$ to denote total spending after step $k$, with $\Sigma t,0$ the pre-pipeline (baseline) spending. We use category-specific subscripts ($\Sigma h$ for hospital, $\Sigma c$ for physician and clinical, $\Sigma p$ for pharmaceuticals, $\Sigma o$ for overhead, $\Sigma e$ for durable medical equipment, $\Sigma d$ for dental) when we operate on individual NHE categories, and aggregate subscripts ($\Sigma service$ for the service-category aggregate of hospital, physician, dental, nursing care,home health, other professional services, and other health, residential and personal care; $\Sigma non\text{-}Other$ for the sum of all categories except 'Other'; $\Sigma included$ and $\Sigma excluded$ as defined in Section 1.8) when we operate on combinations of categories.

#### 1.1 Consolidation of pharmaceutical spending and parallel federal systems

Given that a universal Medicare program would be implemented independently of the Department of Defense (DOD/TRICARE), the Department of Veterans Affairs (VA), and the Indian Health Service (IHS), we separate spending attributable to these entities from the corresponding NHE categories and consolidate them into a single "parallel federal system" category that we exclude from all subsequent pipeline steps.

We also consolidate pharmaceutical spending into a single category that combines retail pharmaceuticals already reported in NHE Table 19 with non-retail pharmaceuticals,[1] that is, drugs administered as a component of service provision in hospital, clinic, nursing home, and home health settings. We estimate non-retail pharmaceutical spending by scaling the Altarum 2022 projection ($720.1 billion, combining retail and non-retail) to the 2024 NHE retail-pharma total and taking the difference.[3] The reallocated non-retail total of $299.06 billion is distributed across hospital (26.3%), physician and clinical (58.3%), nursing care (11.5%), and home health (3.9%) settings, using the Altarum sectoral allocation.[4]

This step changes the categorical composition of spending but does not change its total. Therefore $\Sigma_{t,1} = \Sigma_{t,0} = \$5,278.55$ *billion*.

#### 1.2 Uncompensated hospitalization care

Hospitals incur uncompensated care costs each year when they treat uninsured or underinsured patients who cannot pay their bills. We estimate uncompensated hospital care at $51.69 billion nationally in 2024, derived from the American Hospital Association's 2020 estimate of $42.67 billion,[5] inflated to 2024 dollars using the Bureau of Labor Statistics Consumer Price Index.[6,7] Under universal coverage, all healthcare delivered would be reimbursed; uncompensated care is therefore added back to hospital spending:

$$\Sigma_{h,2} = \Sigma_{h,1} + \$51.69 \text{ billion}$$

#### 1.3 Avoidable emergency room visits and hospitalizations

Avoidable emergency room visits and hospitalizations can be averted through improved access to primary and preventive care under universal coverage. We estimate national savings of $9.99 billion in emergency room expenses and $90.04 billion in avoidable hospitalization costs in 2024.[2,6] We apply these savings to hospital spending:

$$\Sigma_{h,3} = \Sigma_{h,2} - (\$9.99 + \$90.04) \text{ billion}$$

#### 1.4 Reimbursement rate harmonization

A universal Medicare program would establish reimbursement rates for hospital and physician/clinical services comparable to those currently paid by Medicare. The population is currently covered by a mix of private insurance, Medicaid, CHIP, and existing Medicare; standardizing it toward Medicare rates therefore produces a net reduction in payments to providers, since Medicare rates are well below private rates, partially offset by an increase relative to Medicaid, since Medicare rates exceed Medicaid rates. We derive the four rate multipliers for this calculation: Medicare-to-private and Medicare-to-Medicaid ratios for both hospital services and physician/clinical services.

#### Hospital rates: Medicare vs. private

We derive the Medicare-to-private hospital rate ratio from the 2025 Milliman *Commercial Reimbursement Benchmarking* analysis,[8] which reports volume-weighted commercial-to-Medicare rate ratios of 209% for inpatient services, 263% for outpatient services, and 148% for professional services, with corresponding spending volumes of $63,039 million, $107,505 million, and $110,799 million, respectively.

Given these volumes are commercial spending, the implied Medicare-rate spending for each service line is the commercial volume divided by its commercial-to-Medicare ratio, Medicare base = commercial volume / (commercial/Medicare ratio):

*A = $63,039 / 2.09 = $30,162 million (inpatient Medicare base)*

*B = $107,505 / 2.63 = $40,876 million (outpatient Medicare base)*

*C = $110,799 / 1.48 = $74,864 million (professional Medicare base)*

Because professional services are reported separately in the NHE as physician and clinical spending, we exclude the professional volume from the hospital-rate calculation:

*($63,039 + $107,505) / (A + B) = $170,544 / $71,038 = 2.40*

We therefore set the Medicare-to-private hospital rate multiplier at $r_h^{priv} = 1 / 2.40 = 0.42$.

#### Hospital rates: Medicare vs. Medicaid

We derive the Medicare-to-Medicaid hospital rate ratio from the Medicaid and CHIP Payment and Access Commission's (MACPAC) analysis of state Medicaid hospital payment.[9,10] MACPAC reports that Medicaid fee-for-service inpatient hospital base payments were 22% below comparable Medicare rates, implying *Medicaid hospital rate = 0.78 × Medicare hospital rate*, and therefore $r_h^{Mcd} = 1 / 0.78 \approx 1.28$.

#### Physician and clinical rates: Medicare vs. private

For physician and clinical services, we use the same Milliman 2025 analysis.[8] Commercial professional rates are reported at 148% of Medicare (i.e. commercial = 1.48 × Medicare), implying $r_c^{priv} = 1 / 1.48 \approx 0.68$.

#### Physician and clinical rates: Medicare vs. Medicaid

We derive the Medicare-to-Medicaid clinical rate ratio from the reported Medicaid physician fee index in 2024,[11] which implies that Medicaid physician fees were approximately 71% of Medicare physician fees. Therefore *Medicaid physician rate = 0.71 × Medicare physician rate*, and $r_c^{Mcd} = 1 / 0.71 = 1.41$.

#### Application

We apply the four rate multipliers ($r_h^{priv} = 0.42$, $r_h^{Mcd} = 1.28$, $r_c^{priv} = 0.68$, $r_c^{Mcd} = 1.41$) to the post-step-3 spending, weighted by the national payer mix. The resulting category-level reduction factors $Ih$ and $Ic$ are applied to the post-step-3 subtotals:

$$\Sigma h,4 = \Sigma h,3 \times (1 - Ih)$$

$$\Sigma c,4 = \Sigma c,3 \times (1 - Ic)$$

The combined effect of this step is a reduction of $295.56 billion.

### 1.5 Pharmaceutical price negotiation

The US has long prohibited Medicare from negotiating pharmaceutical prices directly with manufacturers, resulting in US drug prices substantially above those in peer countries. The Inflation Reduction Act of 2022 partially relaxed this prohibition for a subset of Medicare-covered drugs, but the bulk of US pharmaceutical spending continues to occur at prices well above international reference levels.[12]

We apply a 51% reduction to pharmaceutical spending, reflecting the savings achievable if US prices were brought to an international reference price derived as a weighted average of prices paid in other high-income countries.[13] We denote the pharmaceutical price reduction as $In = 0.51$ and apply it as:

$$\Sigma p,5 = \Sigma p,4 \times (1 - In)$$

### 1.6 Administrative overhead reduction

Insurance overhead in the US varies widely across payer types, from approximately 2.3% under traditional Medicare[14] to substantially higher rates in commercial insurance, where administrative costs include marketing, claims processing, profit margins, and provider-side billing complexity. Universal Medicare would replace the existing administrative apparatus with a Medicare-equivalent administrative structure.

We re-base overhead to 2.3% of the post-step-5 service, pharmaceutical, and equipment subtotals by directly rebasing the current overhead to the target rate:

$$Io = 0.023$$

$\Sigma o,6 = Io \times (\Sigma service,5 + \Sigma p,5 + \Sigma e,5)$

#### 1.7 Fraud detection

A unified single-payer billing system facilitates fraud detection through consolidated claims data and consistent provider auditing. We apply an 8% reduction across non-Other categories, that is, excluding non-durable medical products, investment, public health, and the parallel federal systems category, based on estimates from a national health insurance fraud detection analysis from Taiwan[15]:

$If = 0.08$

$\Sigma t,7 = (\Sigma t,6 - \Sigma non\text{-}Other,6) + (1 - If) \times \Sigma non\text{-}Other,6$

#### 1.8 Coverage-induced utilization uplift

Given that universal coverage would provide adequate insurance to those currently uninsured or underinsured, we assume that it raises healthcare utilization among the previously uninsured and underinsured toward the levels observed among the adequately insured. We model this uplift through a factor $F$ that combines the shares of currently uninsured and underinsured individuals with their per-capita spending ratios relative to the adequately insured.

We define $F$ as:

$F = [(1 - Pw - Pu) + Pw \times Iw + Pu \times Iu] / [1 - Pw \times (1 - Sw) - Pu \times (1 - Su)]$

where $Pw = 0.081$ and $Pu = 0.138$ are the currently uninsured[16] and underinsured[17] population shares; $Sw = 0.501$ and $Su = 0.860$ are the per-capita spending ratios of the uninsured[18] and underinsured[2] relative to the adequately insured; and $Iw = Iu = 1$ are the post-transition utilization parameters, indicating full parity with the adequately insured under universal coverage.

We apply $F$ to all spending categories except dental, non-durable medical products, investment, public health, and the parallel federal systems category, that is, to all categories of type Service, Pharma, Equipment, and Overhead other than Dental:

$\Sigma t,8 = (\Sigma t,7 - \Sigma included,7) + F \times \Sigma included,7 = \Sigma excluded + F \times \Sigma included,7$

Dental is excluded from this step because dental utilization is modeled separately in Section 1.9, using dental-specific coverage and utilization data. The combined effect of this step is an increase of $197.68 billion.

#### 1.9 Dental coverage

Universal coverage would extend dental benefits to the entire population. We model this step as a change in dental utilization, and apply it to the post-step-8 dental subtotal, $\Sigma d,8$ = \$170.74 billion. Because Step 8 excludes dental, $\Sigma d,8 = \Sigma d,7$, and the step is idempotent with respect to re-execution.

##### Utilization

Dental care is currently rationed by cost to a greater degree than medical care. We use data from the ADA Health Policy Institute's report on dental care use, insurance coverage and cost barriers,[19] which reports, for adults aged 19–64 years in 2022, the proportion with a dental visit in the past year by insurance status and the distribution of dental coverage (**Table S1**).

**Table S1.** Dental visit rates and coverage shares, adults 19–64 years, 2022.

| **Dental coverage** | **Visit rate (*V*)** | **Coverage share (*w*)** |
|---|---|---|
| Private | 0.531 | 0.624 |
| Public | 0.239 | 0.158 |
| Uninsured | 0.152 | 0.218 |

The current population dental visit rate is the coverage-weighted average:

$U_{current} = 0.624 \times 0.531 + 0.158 \times 0.239 + 0.218 \times 0.152 = 0.402$

which reproduces the all-adult visit rate of 40.2% reported in the same source, providing an internal check on the weighting.

Under universal dental coverage, we assume that average utilization rises to the rate currently observed among the privately insured. This gives a utilization multiplier

$m = 0.531 / 0.402 = 1.32$

which we apply to the dental subtotal:

$\Sigma d,9 = \Sigma d,8 \times m = 225.39$ billion

Universal dental coverage therefore raises dental spending by US\$54.65 billion, from US\$170.74 billion to US\$225.39 billion.

**Table S2.** Stepwise transformation of 2024 national health expenditure ($ billions).

| Step | Adjustment | Change | Cumulative |
|---|---|---|---|
| | Baseline (2024 NHE) | | 5278.6 |
| 1 | Parallel federal systems and pharmaceutical consolidation | 0.0 | 5278.6 |
| 2 | Uncompensated hospitalization care | +51.7 | 5330.2 |
| 3 | Avoidable emergency room visits and hospitalizations | −100.0 | 5230.2 |
| 4 | Reimbursement rate harmonization | −295.6 | 4934.7 |
| 5 | Pharmaceutical price negotiation | −377.5 | 4557.1 |
| 6 | Administrative overhead reduction | −286.3 | 4270.8 |
| 7 | Fraud detection | −285.7 | 3985.1 |
| 8 | Coverage-induced utilization uplift | +197.7 | 4182.7 |
| 9 | Dental coverage | +54.7 | 4237.4 |
| | **System-wide saving** | | 1041.2 |

#### 1.10 Sensitivity analyses

We assess the sensitivity of the system-wide saving to the two parameters for which the underlying evidence is least direct: the pharmaceutical price reduction ($l$_n) and the fraud reduction ($l$_f) (**Table S3**). For pharmaceutical prices, we consider two alternatives to the base case: a reduction of 23.1% consistent with the prices negotiated by the Department of Veterans

Affairs, derived from the same source as our international reference price[13] (US prescription drug spending of US$360.3 billion falling to US$277.1 billion), and no reduction at all. For fraud, we remove the reduction entirely. Under every variation, the system-wide saving remains substantial, at no less than US$663.3 billion (12.6% of the current national health expenditure).

**Table S3**: Sensitivity of the system-wide saving to principal expenditure parameters.

| Scenario | System-wide saving ($B) | % of baseline NHE | Difference from base ($B) |
|---|---|---|---|
| Base case | 1,041.2 | 19.7% | — |
| Pharmaceutical pricing: VA-equivalent | 834.4 | 15.8% | −206.8 |
| Pharmaceutical pricing: no reduction | 663.3 | 12.6% | −377.9 |
| Fraud reduction switched off | 733.5 | 13.9% | −307.7 |

### 2. Mortality model

We estimate the annual mortality effect of universal coverage using an attributable-fraction framework applied to 2024 deaths.

#### 2.1 Framework

Within each age group $a$, we treat the observed deaths $D_a$ as arising from a mixture of three insurance strata whose mortality rates differ: the adequately insured, the underinsured, and the uninsured. Let $u_a$ denote the uninsured share of the population in age group $a$, $t_a$ the underinsured share, and $(1 - u_a - t_a)$ the adequately insured share. Let $\lambda$ denote the mortality hazard ratio of the uninsured relative to the adequately insured, and $\gamma$ the corresponding hazard ratio for the underinsured.

Under this mixture, observed deaths relate to the deaths that would occur if the entire age group were adequately insured, $D_a^*$, by:

$$D_a = D_a^* \times [1 + (\lambda - 1)\, u_a + (\gamma - 1)\, t_a]$$

Rearranging gives the counterfactual deaths under universal adequate insurance:

$$D_a^* = D_a / [1 + (\lambda - 1)\, u_a + (\gamma - 1)\, t_a]$$

and the annual lives saved:

$L = \Sigma a (D\_a - D\_a^*)$

### 2.2 Uninsured hazard ratio

We assign the uninsured a mortality hazard ratio of $\lambda = 1.40$ relative to the adequately insured, following cohort estimates of excess mortality among uninsured adults.[20]

### 2.3 Underinsured hazard ratio

Direct estimates of excess mortality among the underinsured are not available, because underinsurance is defined by out-of-pocket burden relative to income rather than by coverage status, and is not recorded in mortality-linked cohorts. We therefore derive $\gamma$ by interpolation.

We assume that the excess mortality of a stratum, $(HR - 1)$, scales with that stratum's position between the adequately insured and the uninsured on an axis measuring the degree to which cost impedes care:

$\gamma = 1 + (\lambda - 1) \times w$, where $w = (\text{underinsured deficit}) / (\text{uninsured deficit})$

#### Baseline: cost-related forgone care

For our baseline, we take $w$ from the prevalence of cost-related barriers to care, the mechanism through which inadequate coverage is understood to raise mortality. The Commonwealth Fund Biennial Health Insurance Survey reports the share of adults reporting at least one of four cost-related access problems in the past year, by insurance status: 36% of adults insured all year and not underinsured, 57% of the underinsured, and 70% of those uninsured at any point.[17] This source uses the same definition of underinsurance as the population share applied in Section 2.4. Therefore:

$w = (0.57 - 0.36) / (0.70 - 0.36) = 0.618$

$\gamma = 1 + (1.40 - 1) \times 0.618 = 1.25$

### 2.4 Population and mortality inputs

Population and uninsured counts by age are taken from the American Community Survey,[16] which reports the civilian noninstitutionalized population. The total uninsured count of 27,479,253 reconciles exactly with the newly insured population used in the expenditure model. Deaths by age are taken from national vital statistics for 2024 (Table S4).[21]

Underinsurance is assigned to adults aged 19–64 years only, and set to zero at ages 0–18 and 65 years and over, where near-universal public coverage through CHIP, Medicaid, and Medicare makes underinsurance, as defined here, uncommon. Within the 19–64 range, we allocate a total of $0.23 \times 197{,}947{,}158 = 45{,}527{,}846$ underinsured individuals across age bands in proportion to the uninsured count in each band, capped in each band at the insured population. This allocation assumes that underinsurance and uninsurance share an age distribution, which reflects their common association with lower income and with employment in sectors that do not offer comprehensive coverage.

**Table S4.** Population, coverage, and mortality inputs by age group, 2024.

| Age group | Population | Uninsured | *u* | *t* | Deaths |
|---|---|---|---|---|---|
| <25 | 107,824,853 | 8,900,577 | 0.0825 | 0.0804 | 39,736 |
| 25–34 | 40,840,056 | 5,764,385 | 0.1411 | 0.2876 | 57,827 |
| 35–44 | 45,074,641 | 5,270,606 | 0.1169 | 0.2383 | 97,430 |
| 45–54 | 40,345,654 | 3,988,967 | 0.0989 | 0.2015 | 157,770 |
| 55–64 | 41,292,992 | 3,065,197 | 0.0742 | 0.1513 | 358,196 |
| 65+ | 59,812,326 | 489,521 | 0.0082 | 0.0000 | 2,341,600 |
| **Total** | **335,190,522** | **27,479,253** | | | **3,052,559** |

### 2.5 Results

Applying $\lambda = 1.40$ and $\gamma = 1.25$ gives 62,863 lives saved annually under universal adequate coverage (**Table S5**).

**Table S5.** Lives saved by age group, $\lambda = 1.40$, $\gamma = 1.25$.

| Age group | Deaths observed | Deaths under universal adequate insurance | Lives saved |
|---|---|---|---|
| <25 | 39,736 | 36,126 | 3,610 |
| 25–34 | 57,827 | 51,248 | 6,579 |
| 35–44 | 97,430 | 88,065 | 9,365 |
| 45–54 | 157,770 | 144,754 | 13,016 |
| 55–64 | 358,196 | 335,544 | 22,652 |
| 65+ | 2,341,600 | 2,333,959 | 7,641 |

| **Age group** | **Deaths observed** | **Deaths under universal adequate insurance** | **Lives saved** |
|---|---|---|---|
| **Total** | **3,052,559** | **2,989,696** | **62,863** |

**Table S6.** Sensitivity of lives saved to the underinsured hazard ratio $\gamma$, with $\lambda$ fixed at 1.40.

| $\gamma$ | **Lives saved (all ages)** | **Note** |
|---|---|---|
| 1.00 | 33,232 | No excess mortality attributed to underinsurance |
| 1.10 | 45,422 | |
| 1.20 | 57,158 | |
| **1.25** | **62,863** | **Baseline, cost-related forgone care basis** |
| 1.30 | 68,464 | |
| 1.40 | 79,367 | |

6 Consumer Price Index. U.S. Bureau of Labor Statistics. https://www.bls.gov/cpi/.

7 $42.67 in 2020 → 2024. Official Data. 2026; published online April 30. https://www.in2013dollars.com/us/inflation/2020?amount=42.67 (accessed May 26, 2026).

8 Commercial reimbursement benchmarking 2025: Commercial payment rates for medical services as a percentage of Medicare fee-for-service rates. 2025; published online June 24. https://www.milliman.com/en/insight/commercial-reimbursement-benchmarking-medicare-ffs-rates-2025 (accessed May 29, 2026).

9 Medicaid Hospital Payment: A Comparison across States and to Medicare. MACPAC. 2017; published online April. https://www.macpac.gov/wp-content/uploads/2017/04/Medicaid-Hospital-Payment-A-Comparison-across-States-and-to-Medicare.pdf (accessed May 29, 2026).

10 Mann C, Striar A. How differences in medicaid, medicare, and commercial health insurance payment rates impact access, health equity, and cost. 2022. DOI:10.26099/C71G-3225.

11 Skopec L, Pugazhendhi A, Zuckerman S. Updated Medicaid-to-Medicare fee index: Medicaid physician fees still lag behind Medicare physician fees. *Health Aff (Millwood)* 2025; **44**: 531–8.

12 Inflation Reduction Act of 2022. https://www.irs.gov/inflation-reduction-act-of-2022 (accessed May 29, 2026).

13 Ye Y, Pandey A, Fitzpatrick MC, *et al.* Estimating US savings on outpatient prescription pharmaceuticals from international reference pricing. *Proc Natl Acad Sci U S A* 2026; **123**: e2520871122.

14 A Fiscal Analysis of Elizabeth Warren's 'Medicare for All' Plan. The Foundation for Research on Equal Opportunity. https://freopp.org/whitepapers/a-fiscal-analysis-of-elizabeth-warrens-health-care-plan/ (accessed May 29, 2026).

15 Lu J-FR, Hsiao WC. Does universal health insurance make health care unaffordable? Lessons from Taiwan. *Health Aff (Millwood)* 2003; **22**: 77–88.

16 U.S. Census Bureau. Explore Census Data. https://data.census.gov/table/ACSST1Y2024.S2701 (accessed July 16, 2026).

17 Collins SR, Gupta A. The state of health insurance coverage in the U.s.: Findings from the commonwealth fund 2024 biennial health insurance survey. 2024. DOI:10.26099/BYCE-QC28.

18 Coughlin TA, Holahan J, Caswell K, McGrath M. Uncompensated Care for the Uninsured in 2013: A Detailed Examination. KFF. 2014; published online May 30. https://www.kff.org/affordable-care-act/uncompensated-care-for-the-uninsured-in-2013-a-detailed-examination/ (accessed June 1, 2026).

19 National Trends in Dental Care Use, Dental Insurance Coverage, and Cost Barriers. Health Policy Institute. 2024. https://www.ada.org/-/media/project/ada-organization/ada/ada-org/files/resources/research/hpi/national_trends_dental_use_benefits_barriers_2024.pdf (accessed July 16, 2026).

20 Wilper AP, Woolhandler S, Lasser KE, McCormick D, Bor DH, Himmelstein DU. Health insurance and mortality in US adults. *Am J Public Health* 2009; **99**: 2289–95.

21 Xu J, Murphy SL, Kochanek KD, Arias E. Mortality in the United States, 2024. *NCHS Data Brief* 2026; published online Jan. DOI:10.15620/cdc/174641.
